# High power transient 12-29Hz beta event features as early biomarkers of Alzheimer’s Disease conversion: a MEG study

**DOI:** 10.1101/2024.09.13.24313611

**Authors:** Danylyna Shpakivska-Bilan, Gianluca Susi, David W Zhou, Jesus Cabrera, Blanca P Carvajal, Ernesto Pereda, Maria Eugenia Lopez, Ricardo Bruña, Fernando Maestu, Stephanie R Jones

## Abstract

A typical pattern observed in M/EEG recordings of Mild Cognitive Impairment (MCI) patients progressing to Alzheimer’s disease is a continuous slowing of brain oscillatory activity. Definitions of oscillatory slowing are imprecise, as they average across time and frequency bands, masking the finer structure in the signal and potential reliable biomarkers of the disease.

Recent studies show that high averaged band power can result from transient increases in power, termed ‘events’ or ‘bursts’. To better understand MEG oscillatory slowing in AD progression, we analyzed features of high-power oscillatory events and their relationship to cognitive decline. MEG resting-state oscillations were registered in age-matched patients with MCI who later convert (CONV, N=41) or do not convert (NOCONV, N=44) to AD, in a period of 2.5 years. To distinguish future CONV from NOCONV, we characterised the rate, duration, frequency span and power of transient high-power events in the alpha and beta band in anterior cingulate (ACC) and precuneus (PC).

Results revealed event-like patterns in resting-state power in both the alpha and beta-bands, however only beta-band features were predictive of conversion to AD, particularly in PC. Specifically, compared to NOCONV, CONV had a lower number of beta events, along with lower power events and a trend toward shorter duration events in PC (*p <* 0.05). Beta event durations were also significantly shorter in ACC (*p <* 0.01). Further, this reduced expression of beta events in CONV predicted lower values of mean relative beta power, increased probability of AD conversion, and poorer cognitive performance.

Our work paves the way for reinterpreting M/EEG slowing and examining beta event features as a new biomarker along the AD continuum, and a potential link to theories of inhibitory cognitive control in neurodegeneration. These results may bring us closer to understanding the neural mechanisms of the disease that help guide new therapies.

## 1 Introduction

According to the World Health Organization 2017 Alzheimer’s disease (AD), a leading cause of disability and dependency among older individuals worldwide, is expected to affect 130 million people by 2050. Despite intensive research efforts, disease-modifying human therapies are still lacking, since the link between amyloid-induced cellular damage and cognitive decline is incomplete (Maestú et al., 2021).

Magnetoencephalography (MEG) has been an valuable technique to fill this gap, as it can directly capture human neuronal processes, associated with the disease and cognition, with high temporal resolution (da Silva, 2013; Maestú et al., 2021). A typical pattern observed in M/EEG recordings of AD patients is a progressive slowing of brain oscillatory activity (Dauwels et al., 2011; Hsiao et al., 2013; Ishii et al., 2017; Jeong, 2004), typically characterised by an increase in low-frequency delta (0.5 – 4 Hz) and theta rhythms (4-7 Hz), along with a decrease in higher frequency bands, alpha (8–12 Hz) and beta (12–30 Hz) rhythms. This oscillatory slowing initiates in early stages of the disease, such as Mild Cognitive Impairment (MCI) (Babiloni et al., 2004, 2009, 2010; Dauwels et al., 2011; Jelic et al., 2000), and may even manifest before, in the subjective cognitive decline stage (Bruña et al., 2023; Ĺopez-Sanz et al., 2016), progressing from anterior to posterior cortices and particularly in frontal and parietal regions, (Huang et al., 2000; Nakamura et al., 2018), in line with the onset of amyloid accumulation in the fronto-temporal association cortices (Bang et al., 2015; Cho et al., 2016; Wiesman et al., 2022). It has been shown that MCI patients who finally convert to AD exhibit a significant disruption (i.e., decrease in synchronisation (König et al., 2005; Ĺopez-Sanz et al., 2017; Pusil, Dimitriadis, et al., 2019), as stated in the “X” model) between anterior cingulate cortex (ACC) and precuneus (PC), two regions typically associated with high amyloid deposition (Forsberg et al., 2008). As MEG oscillatory slowing accelerates, cognitive decline worsens producing alterations in memory processes and executive functions (Hoshi et al., 2022; Wiesman et al., 2022).

Definitions of oscillatory slowing are imprecise, as they typically rely on methods based on a spectral decomposition followed by averaging across time, frequency bands, and often subjects. Such averaging can mask finer structure in the signal that may provide more reliable biomarkers of the disease and cognitive decline and help connect human biomarkers to the underlying neural mechanisms of the disease including possible connections to hyperexcitability as shown in animal models (Maestú et al., 2021; Stoiljkovic et al., 2018; Zott et al., 2019). In recent years, there has been a shift in spectral M/EEG methods, as many studies have shown that, in non-averaged data, brain oscillations often occur as transient increases in high spectral power, a phenomenon termed oscillatory “bursts” or “events” (Jones, 2016; Lundqvist et al., 2024; van Ede et al., 2018). Quantifying transient changes in spectral activity requires new methods that consider temporal characteristics of spectral activity such as event rate, amplitude, duration, or frequency span (Shin et al., 2017). Such event-based methods have recently been applied in a growing body of M/EEG studies on the brain dynamics of cognitive processes (Kavanaugh et al., 2023, 2024; McKeon et al., 2023; Morris et al., 2023; Quinn et al., 2019; Shin et al., 2017), helping to establish neural correlates of cognitive behavior on a single trial level. Variability in oscillatory event parameters may represent a new set of explainable MEG biomarkers for AD, as it can reflect differences in circuit-level origins and provide insights into the underlying activity patterns and functions (Jones, 2016; Lundqvist et al., 2024; M. A. Sherman et al., 2016).

In this study we applied standard power spectral density (PSD) and event-based analysis methods to resting-state MEG from adults with MCI who later convert (CONV) or do not convert (NOCONV) to AD. Motivated by the findings of the AD continuum model described by (Pusil, López, et al., 2019) (namely the “X” model), we first hypothesized that averaged PSD slowing exhibits divergent effects in features of high-power transient spectral events. Second, we hypothesized that slowing-related effects in spectral event features would be associated to cognitive decline, as measured by a battery of neuropsychological tests in memory and executive functions in the MCI sample. We characterize MEG oscillatory slowing in terms of transient spectral event parameters in a MCI-to-AD longitudinal sample, taking the initial step towards the potential identification of biophysically principled biomarkers.

## 2 Methods

### 2.1 Subject recruitment and neuropsychological assessment

Participants were recruited from Hospital Cĺınico Universitario San Carlos in Madrid, Spain. The study was approved by the Ethics Committee, and all participants provided written informed consent prior to participation. All participants were right-handed native Spanish speakers.

The study sample included 85 subjects diagnosed with mild cognitive impairment (MCI). Initially, participants were screened according to the diagnostic criteria of the National Institute on Aging-Alzheimer’s Association (NIA-AA) (Albert et al., 2011) and underwent a comprehensive neuropsychological assessment as previously described (Ĺopez et al., 2016; Pusil, Dimitriadis, et al., 2019), along with a MEG recording. They were cognitively and clinically followed-up in a temporal interval of 2.5 years and then subdivided in two groups considering the criteria for probable Alzheimer’s disease (McKhann et al., 2011): 41 subjects with mild cognitive impairment who converted to AD (CONV), and 44 subjects with mild cognitive impairment (MCI) which did not convert to AD (NOCONV). Subjects in CONV and NOCONV group were matched by age, sex, and education years, as it is reported in reported in Table 1.

**Table 1:** Mean ± SD values of the demographic and neuropsychological characteristics for AD converters (CONV) and NOCONV groups. Non significant p-values are marked as n.s.

| Descriptives | NO CONV Mean ± SD | CONV Mean ± SD | p value |
| --- | --- | --- | --- |
| Age | 75.25 ± 5.36 | 75.39 ± 4.99 | t (83), p = n.s. |
| Sex | N= 44; 19 (m), 25 (f) | N= 41; 17 (m), 24 (f) | Xi (1), p = n.s. |
| Education years | 9.89 ± 5.39 | 8.32 ± 4.30 | t (71), p = n.s. |
| Immediate Logic Memory Units | 17.30 ± 12.61 | 10.89 ± 7.41 | t (83), p = .01 |
| Delayed Logic Memory Units | 7.64 ± 9.10 | 2.20 ± 4.08 | t (83), p < .01 |
| Immediate Logic Memory Themes | 10.60 ± 5.03 | 7.70 ± 3.83 | t (83), p < .01 |
| Delayed Logic Memory Themes | 4.86 ± 4.52 | 2.29 ± 2.47 | t (83), p < .01 |
| Inverse Digits | 4.16 ± 1.79 | 4.15 ± 1.28 | t (83), p = n.s. |
| Semantic Fluency | 11.86 ± 3.56 | 11.15 ± 3.60 | t (83), p = n.s. |
| Trail Making Test - B | 211.15 ± 112.72 | 251.34 ± 108.15 | t (83), p = n.s. |
| Boston Naming Test | 46.45 ± 9.77 | 43.80 ± 8.74 | t (83), p = n.s. |
| Clock-Drawing Test | 5.43 ± 2.08 | 5.07 ± 2.26 | t (83), p = n.s. |

The neuropsychological assessment included seven tests: four measures of memory recall, *Immediate Logic Memory Units*, *Delayed Logic Memory Units*, *Immediate Logic Memory Themes* and *Delayed Logic Memory Themes* (Wechsler Memory Scale,WMS-III) (Wechsler, 1997); one measure of working memory, *Inverse Digits* (WMS-III); one measure of cognitive flexibility, *Trail Making Test-B* (TMT-B) (Bowie & Harvey, 2006); two language measures, *Semantic Fluency* (Controlled oral Word Association Test, COWAT) (Benton et al., 1994) and *Boston Naming Test* (BNT) (Kaplan et al., 1983) ; and one global screening measure for cognitive impairment and dementia, the *Clock-Drawing copy test* (Agrell & Dehlin, 1998). Table 1 includes paired t-test and previously reported differences across groups (López-Sanz et al., 2016; Pusil, Dimitriadis, et al., 2019).

### 2.2 Magnetoencephalography data acquisition

The dataset was acquired using a 306-channel (102 magnetometers and 204 gradiometers) Vectorview MEG system (Elekta AB, Stockholm, Sweden) placed inside a magnetically shielded room (Vacuum-Schmelze GmbH, Hanau, Germany) located at the Laboratory of Cognitive and Computational Neuroscience (Madrid, Spain). MEG data consisted of 5 min eyes-closed resting-state recordings in a 60 min session, with a sampling rate of 1000 Hz and an online [0.1 - 330] Hz anti-alias band-pass filter. To allow further analysis, including subject-specific source reconstruction, MEG recordings were complemented by MRI scans acquired within a month after the MEG session, which were recorded at the Hospital Universitario Cĺınico San Carlos (Madrid, Spain) using a 1.5 T General Electric MRI scanner with a high-resolution antenna and a homogenization PURE filter (fast spoiled gradient echo sequence, with parameters: repetition time/echo time/inversion time = 11.2/4.2/450 ms; flip angle = 12°; slice thickness = 1 mm; 256×256 matrix; field of view = 256 mm).

The MEG recordings were preprocessed offline using a temporal-spatial filtering algorithm (tSSS) (Taulu & Hari, 2009) (Maxfilter Software v2.2, correlation limit of 0.9 and correlation window of 10 s) to eliminate magnetic noises and compensate for head movements during the recording. The continuous MEG data were imported into MATLAB (R2017b, Mathworks, Inc.) for pre-processing steps, carried out using the Fieldtrip Toolbox (Oostenveld et al., 2011) (https://www.fieldtriptoolbox.org/). Data were automatically scanned for ocular, muscle and jump artefacts using the Fieldtrip software. Artefacts were then visually confirmed by an MEG expert. The remaining artefact-free data were segmented into 4-s segments (epochs). An independent component analysis-based procedure was used to remove the heart magnetic field artefact. Source reconstruction was performed using minimum norm estimates (Hämäläinen & Ilmoniemi, 1994) with the software Brainstorm (Tadel et al., 2011). Current dipoles were constrained to be perpendicular to the individual’s cortical surface, to model the orientation of macro columns of pyramidal neurons (Tadel et al., 2011). Neural time series were finally averaged within regions of interest (ROI) of the Schaefer 100-17 network atlas (Schaefer et al., 2018). Two regions of interest were extracted for subsequent analysis: the anterior cingulate cortex (as the merge of *left ACC* and *right ACC*, corresponding to *SalV entAttnB PFCmp* 1 area of the mentioned atlas, respectively), and the precuneus (as the merge of *left PC* and *right PC*, corresponding to *ContC pCun* 1 and *DefaultA pCunPCC* 1 area of the mentioned atlas), see Figure 1a. The data was band-pass filtered between 0.5 and 45 Hz (broadband), using FIR filtering.

**Figure 1:**
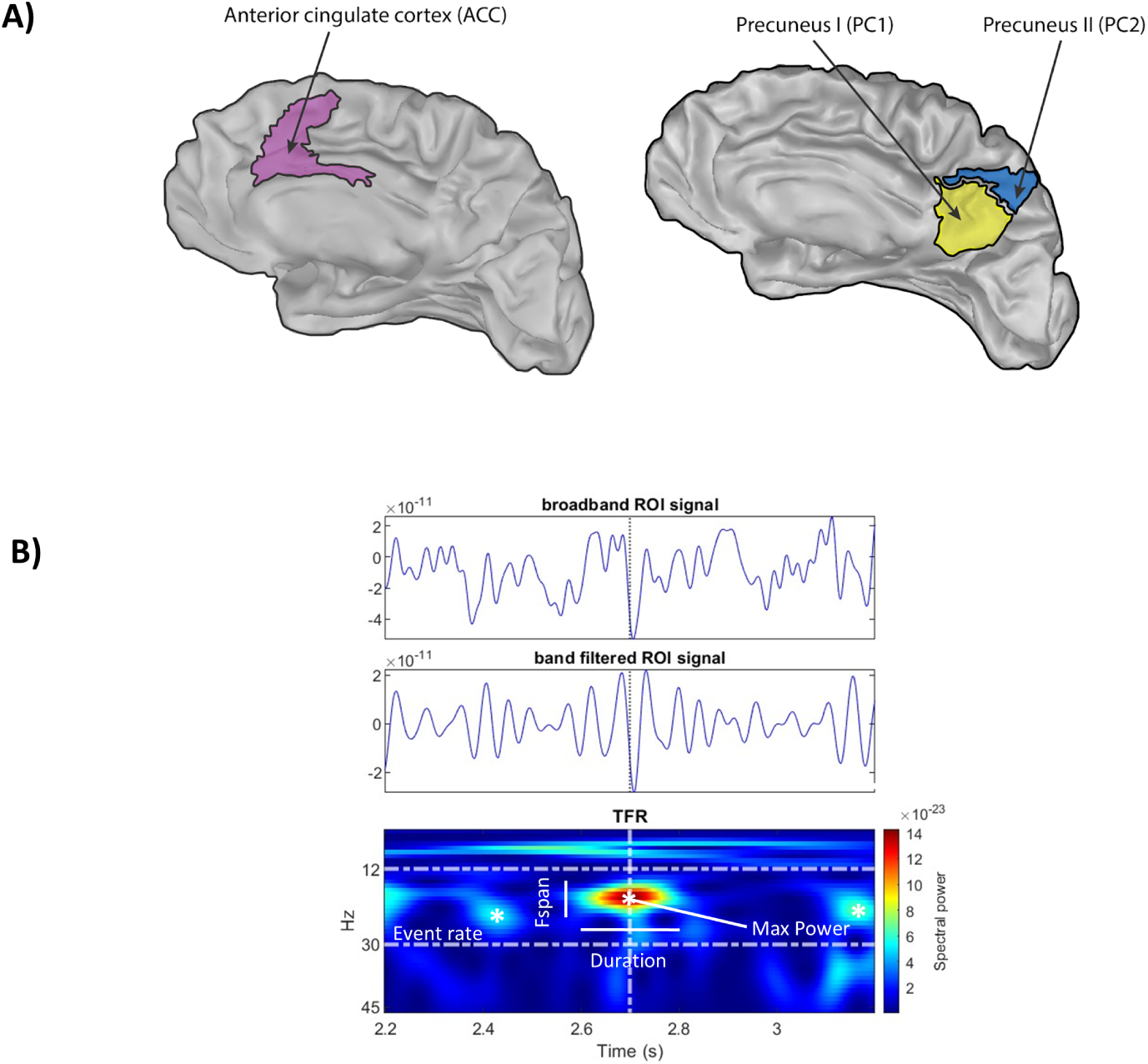
a) Representation of the right side regions of interest using the Schaefer 100-17 network atlas: *SalV entAttnB PFCmp* 1 (corresponding to ACC), *ContC pCun* 1 (corresponding to PC) and *DefaultA pCunPCC* 1 (corresponding to PC); b) Example of resting-state beta activity from left-ACC and detection of transient burst of high-power activity (events), using the frequency-based algorithm of Shin et al. (2017). There are several possible features of such events that could contribute to increased power averaged across time and frequency, including event number (rate), event duration, event frequency span and event power

### 2.3 Power spectral density (PSD)

We computed the power spectral density of each of the ROI time series by using the Welch’s periodogram method (Welch, 1967), with 1s window length and 50% overlap ratio. For each ROI signal, the normalised power was calculated by averaging the power spectral density obtained by each epoch and then normalising the value associated to each frequency by total power over the [1–30] Hz range (Figure 2).

**Figure 2:**
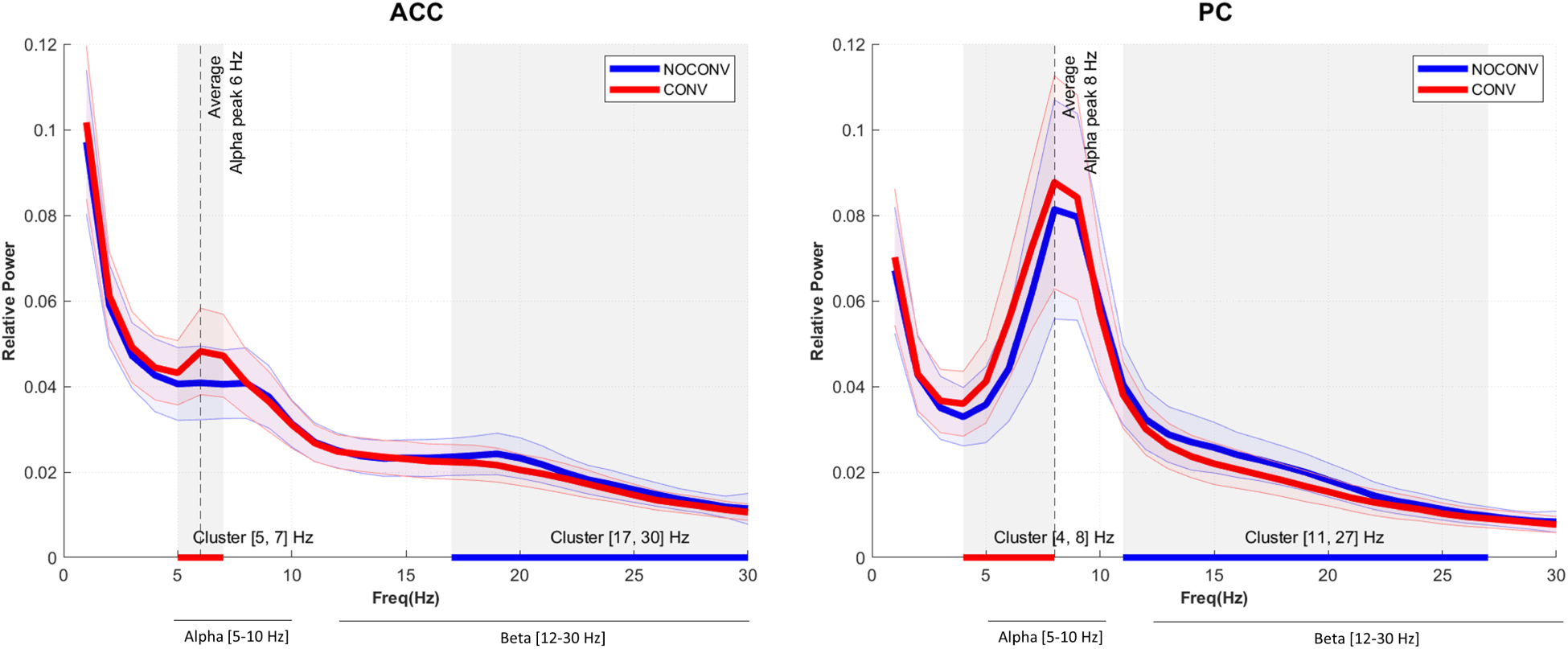
Normalized power spectral density plots (mean ± SEM) for regions of interest in CONV and NOCONV patients, in the range [2-30] Hz. Shaded grey areas represent statistically significant clusters of differences between CONV and NOCONV groups (non-parametric statistical test).

### 2.4 Time-Frequency Representations (TFRs) & Spectral Events Extraction

During resting-state alpha and beta activity, periods of transient high power can be quantified over time in unaveraged data. We use a time-frequency-based algorithm as in Shin et al. (2017) to capture these bursts.

Time-Frequency Representations (TFRs) were calculated using the MATLAB SpectralEvents Toolbox (find method = 1, as in Shin et al. (2017)) (https://github.com/jonescompneurolab/SpectralEvents). Each artefact-free 4-sec epoch was convolved with a 7-cycle Morlet wavelet. For all epochs, TFR across time and across each patient was calculated and finally averaged to obtain a representative TFR for each ROI (PC or ACC) and group (CONV or NOCONV) in the range [2-30] Hz.

The bands of interest reflected the slowing effect typical of MCI (Bruña et al., 2023; Dauwels et al., 2011; López-Sanz et al., 2016) and observed in our sample and were chosen to be [5-10] Hz for alpha and [12-30Hz] for beta. (See also methods section 2.5 *Statistical Analysis*). More specifically, we defined alpha by determining the group averaged peak (GAP) frequency of 8Hz in PC and taking a range of GAP [-3, +2] Hz. This choice provides a more precise capture of alpha oscillations in the ageing population (Tröndle et al., 2023) and reflects the range of significant difference in both PC and ACC across groups in our sample (see results Figure 2 below). The [12-30]Hz beta band, which does not exhibit a clear GAP, was similarly chosen to include the areas of significant difference in our sample.

Spectral Events were detected by first retrieving all local maxima in un-normalized TFR using *imregional-max*. For each subject and ROI, transient high-power events were defined as local maxima above a 6X factor of the median (FOM) threshold within a frequency band of interest, to be consistent with prior studies (Shin et al., 2017). Additionally, we tested the robustness of our analysis with 4X FOM and 8X FOM thresholds (see Supplementary Figure 2 and Supplementary Table 1 & 2). As reported, we had the most significant results for 6X FOM median threshold. The spectral event method applied (namely, find method=1 in the SpectralEvent Toolbox) allows for multiple, overlapping events to occur in a given suprathreshold region and does not guarantee the presence of within-band, suprathreshold activity in any given trial, see Figure 1b.

Importantly, it was noticed that events detected in contralateral medial ROIs (especially in left ACC and right ACC) were in part mirrored across the midline due to the close proximity of neighbouring bilateral dipoles (this effect of spatial smearing in source reconstruction is explained in Supplementary Figure 1). To remove duplicated mirror events when two detected events shared the corresponding contralateral ROI, epoch, time and frequency, we rejected the one with lower amplitude.

For each subject and ROI, spectral events were characterised by 4 key features: event number in a fixed time window (i.e. event rate), duration, frequency span (Fspan) and power (Power FOM) (see Figure 1b). Event number was calculated by counting the number of events in the 4-second period of each epoch. Event power was calculated as the normalised FOM power value at each event maximum. Event duration and frequency span were calculated from the boundaries of the region containing power values greater than half the local maxima power, as the full-width-at-half-maximum in the time and frequency domain, respectively.

### 2.5 Statistical analysis

To determine frequency ranges that represent oscillatory slowing in our cohort, we tested for significant differences in average power spectral density between AD CONV and NOCONV for each selected ROI by performing a non-parametric statistical test (Maris & Oostenveld, 2007) in the frequency domain (see light grey window in Figure 2).

In each frequency-band, we examined the relationship between event features and the averaged PSD using a linear regression analysis over all subjects. The regression *β* coefficients were calculated with a 95% CI and all p-values were corrected for multiple comparisons using the Benjamini–Hochberg (BH) step-up procedure (Benjamini & Hochberg, 1995) with a False Discovery Rate (Q) set at 0.05. Statistically significant p-values after BH correction are reported as ∗*p <* 0.05(*Q* = 0.05).

We also tested for group differences in spectral event features between CONV and NOCONV groups. High power spectral event features were detected in alpha and beta frequency bands for each resting state segment from four ROI’s (left and right ACC and PC, respectively). Event rates were averaged across epochs, and other features (duration, f-span, power) were averaged across all events, and then between hemispheres, within each subject. The final dataset consisted of 16 variables for each subject: four event features (averaged spectral event rate, duration, f-span and power); by two ROI’s (ACC, PC); by two frequency bands (alpha and beta). For each variable, t-tests were used to assess group differences in transient event features. The tests were conducted with a left tail for the beta band and a right tail for the alpha band. This approach was consistent with the observation that the average power spectra in the alpha band are higher for CONV compared to NOCONV, while the effect is reversed in the beta band. We hypothesized that differences in event features align with the directional trend observed in the averaged relative spectral power. All reported p-values were corrected for multiple comparisons across the number of tests applied using the Benjamini–Hochberg (BH) step-up procedure (Benjamini & Hochberg, 1995) with a False Discovery Rate (Q) set at 0.05. Statistically significant p-values after BH correction are reported as ∗ ∗ *p <* 0.05(*Q* = 0.05). Statistical tendencies are reported as significant if ∗*p <* 0.1. We computed effect size of the differences between groups with a robust variant of Cohen’s d (Algina et al., 2005).

For event features where significant difference across groups were found, we examined the relationship between these features and AD conversion and cognitive performance.

To assess the relationship with AD conversion, we fit beta event features from ACC and PC to a logistic regression model (R *glm()* function, *family = binomial* argument) in order to predict each patient’s conversion label (CONV or NOCONV). The model’s output is the probability of the positive class (CONV group). We used a default 0.5 threshold value to transform the probability to a binary class. Thus, a subject is classified as class 1 (CONV) if the predicted probability is greater than or equal to 0.5, and class 0 if the probability is less than 0.5. The odds ratio in logistic regression is calculated by exponentiating the coefficient of the predictor variable. As an example, for predictor variable *Number of events*, if the coefficient is *β* = 0.58 the odds ratio is given by OR = *e*^0.58^ ≈ 1.79. This means that the likelihood of the predicted outcome (AD conversion), is approximately 1.79 times greater than the likelihood of the outcome not occurring (no conversion to AD). Every reduction of 1 unit in *Number of events* increases by 79% the odds of AD conversion (CONV group).

To asses the relationship with cognitive performance, we calculated a Cognitive Performance Index and applied linear regression. Aiming to account for variability across the nine neuropsychological tests used, the Cognitive Performance Index was calculated by applying Principal Components Analysis (PCA) (R *princomp()* function) to the neuropsychological testing dataset. As a first step, the data from each of the nine tests was z-scored to prevent biases due to different test scales. Additionally, the data were transformed so that for each test higher values correspond to better cognitive performance. Then, we extracted the first principal component, which explained at least 55% of variance. The projection of individual data onto the new axes (principal component) represent the Cognitive Performance Index values for each subject, and this projection is achieved through linear transformation using the eigenvectors of the covariance matrix. All eigenvectors had a positive load in the first component. A subject with higher values of this Cognitive Performance Index correspond to better cognitive performance.

## 3 Results

### 3.1 AD Converters (CONV) exhibit oscillatory slowing in average PSD

We confirm a slowing effect in power spectral density in CONV subjects for all ROIs of interest in ACC and PC, as observed in prior studies (Pusil, Dimitriadis, et al., 2019). In averaged PSD, the CONV group shows a statistically significant cluster of decreased relative power in beta frequency band compared to NOCONV (Figure 2, 17-30Hz ACC and 11-27Hz PC, blue underlines) and increased relative power in lower frequency bands (theta, low-alpha) (Figure 2, 5-7Hz ACC and 4-8Hz PC, red underlines). Based on our data, for all subsequent analyses, we utilized a [5-10] Hz alpha bands and [12-30] Hz beta-band (see Methods section 2.4 for further details).

### 3.2 Resting-state alpha and beta emerge as transient high-power events

The PSD plots of Figure 2 rely on Fourier analysis performed on averaged epochs of brain activity from the two different groups. As described previously, differences in averaged power across patient groups could emerge from several features in transient of high-power activity (i.e., *events*) in the unaveraged data.

Visual inspection of time-frequency representations in Figure 3 shows that seemingly continuous high average power in the alpha and beta bands (top panels) is the result of the accumulation of transient high power activity (’events’) across epochs of the unaveraged resting-state data (Figure 3 middle and bottom panels) for both groups.

**Figure 3:**
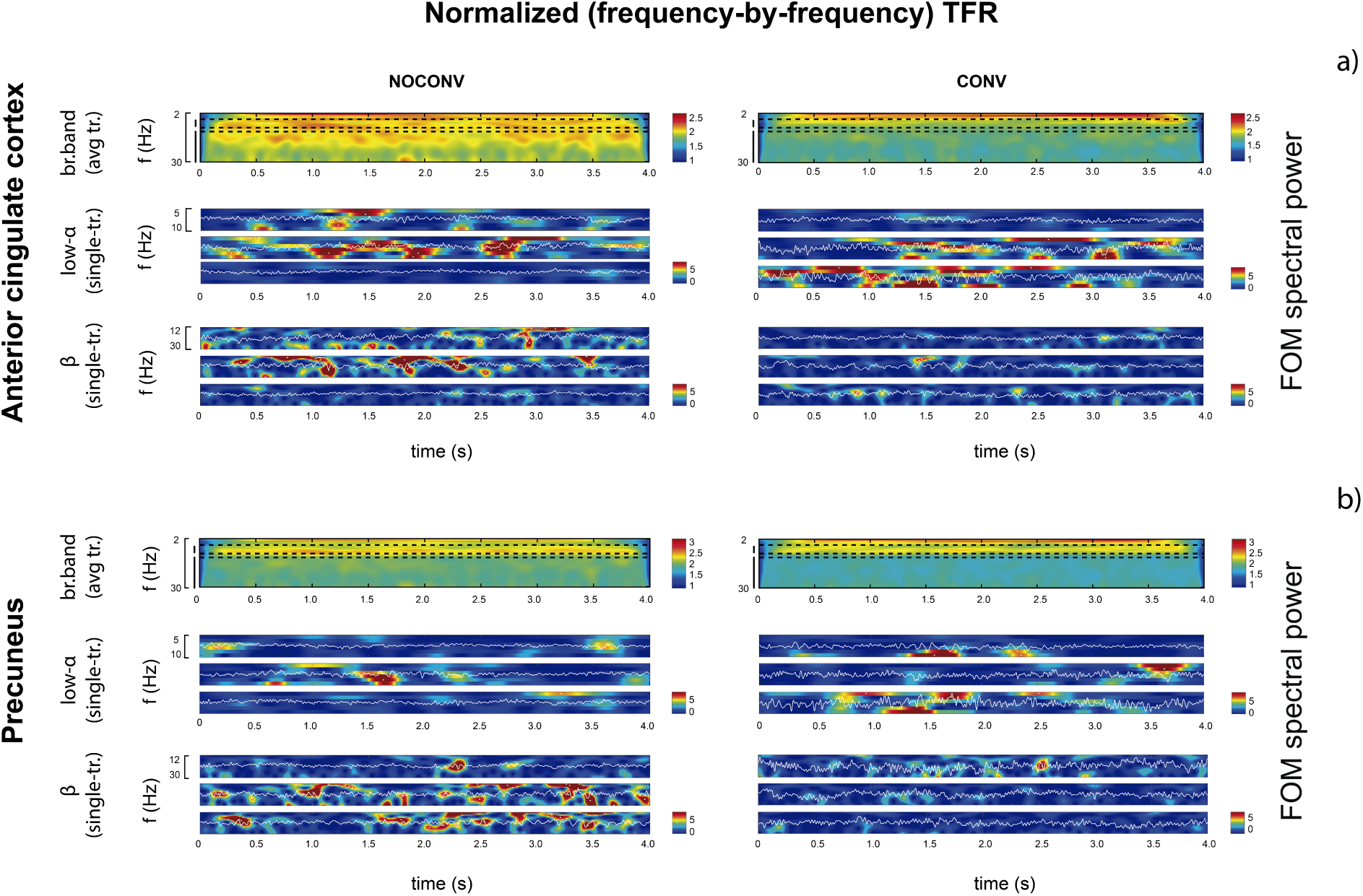
Time-Frequency representations of spectral events in PC (a), and ACC (b) for non-converters (NOCONV, left) and converters (CONV, right) groups. For each case, we show the representative averaged broadband TFR (top), as well as three TFR related to randomly chosen trials for low-alpha (middle), and beta (bottom) band.

As such, higher averaged power could be due to increased expression in several features, including a higher number of events (rate), longer duration events, increased frequency spans and or increased power of the event. To assess if these features contributed to averaged power in our sample, we performed a regression analysis between each feature and averaged alpha and beta power (Figure 4). Our results show a strong correlation with averaged power and each event features in the beta band (number: *β* = .28, *R*^2^ = .069, *p* − *value* = .0088; duration: *β* = .45, *R*^2^ = .192, *p* − *value < .*0001; max power: *β* = .41, *R*^2^ = .156, *p* − *value* = .0001, frequency span: *β* = −.29, *R*^2^ = .07, *p* − *value* = .0075). While similar relationships occurred between alpha event features and averaged alpha power, only alpha event frequency span significantly correlated with averaged alpha power after correction for multiple comparisons and the effect was weaker than in the beta band (frequency span: *β* = −.25, *R*^2^ = .05, *p* − *value* = .025).

**Figure 4:**
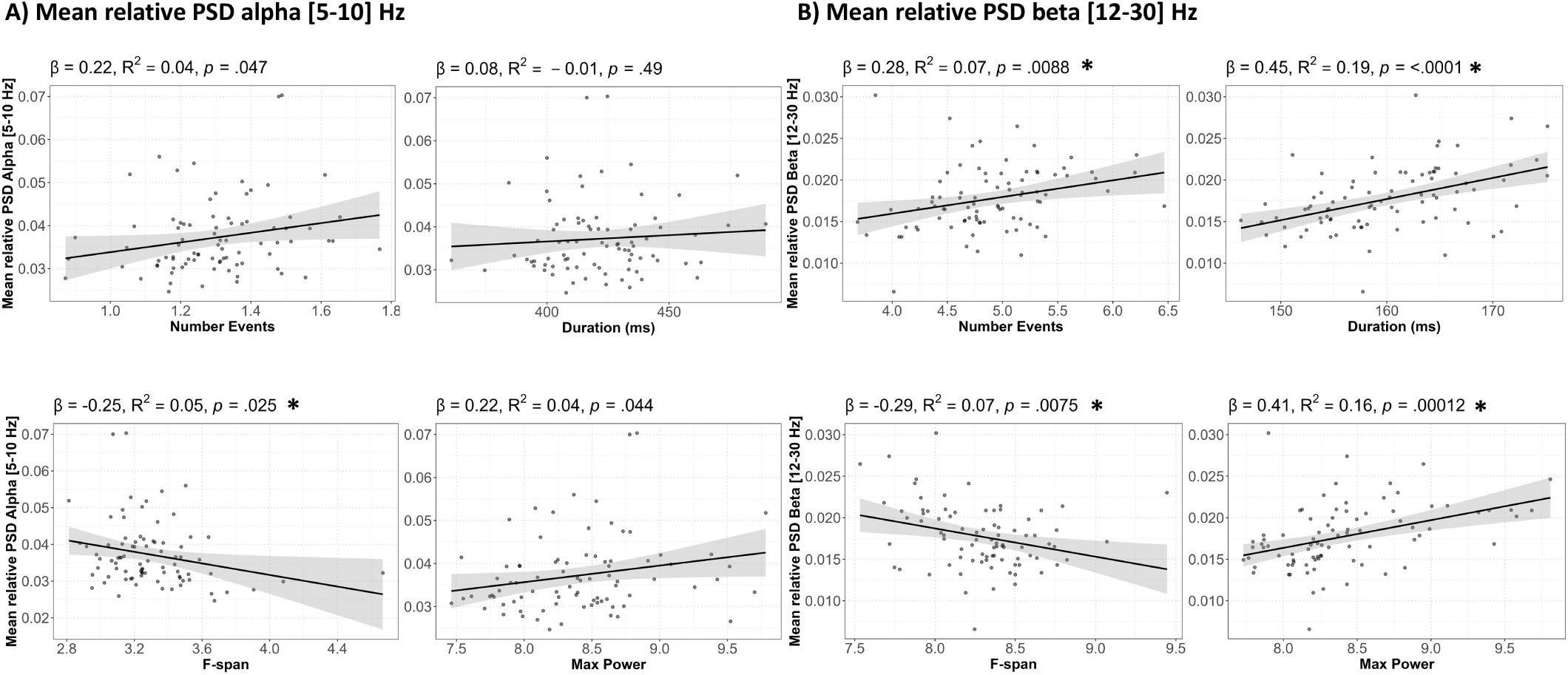
Diminished expression of event features in ACC and PC is associated with reduced mean power spectral density (PSD). A) Regression analysis between alpha [5-10] Hz spectral events and averaged alpha power. B) Regression analysis between beta [12-30] Hz spectral events and averaged beta power. Statistically significant p-values after BH correction are reported as ∗*p <* 0.05(*Q* = 0.05).

### 3.3 Reduced beta event features as predictive biomarkers of AD conversion and cognitive decline

Given the event like nature of alpha and beta, we next tested the hypothesis that the slowing observed differences across groups in averaged PSD power in Figure 2 was due to across group differences in event features, namely rate, power, duration and/or frequency-span. We found effects only in the beta band that were more prominent in PC, such that CONV had a lower mean rate of beta events, lower power events, and a trend to shorter duration beta events in PC. Shorter duration beta events were also present in ACC in the CONV group (Figure 5 and Table 2; rate in PC *p* − *value_BH_*= .003, power in PC *p* − *value_BH_*= .002, duration in PC *p* − *value_BH_*= .051; duration in ACC *p* − *value_BH_*= .01, see also Supplementary Figure 2 and Figure 3 for further summary statistics). Notably, the effect size for differences between CONV and NOCONV in these features are of medium size (*Cohen d >* 0.4), see Table 2).

**Figure 5:**
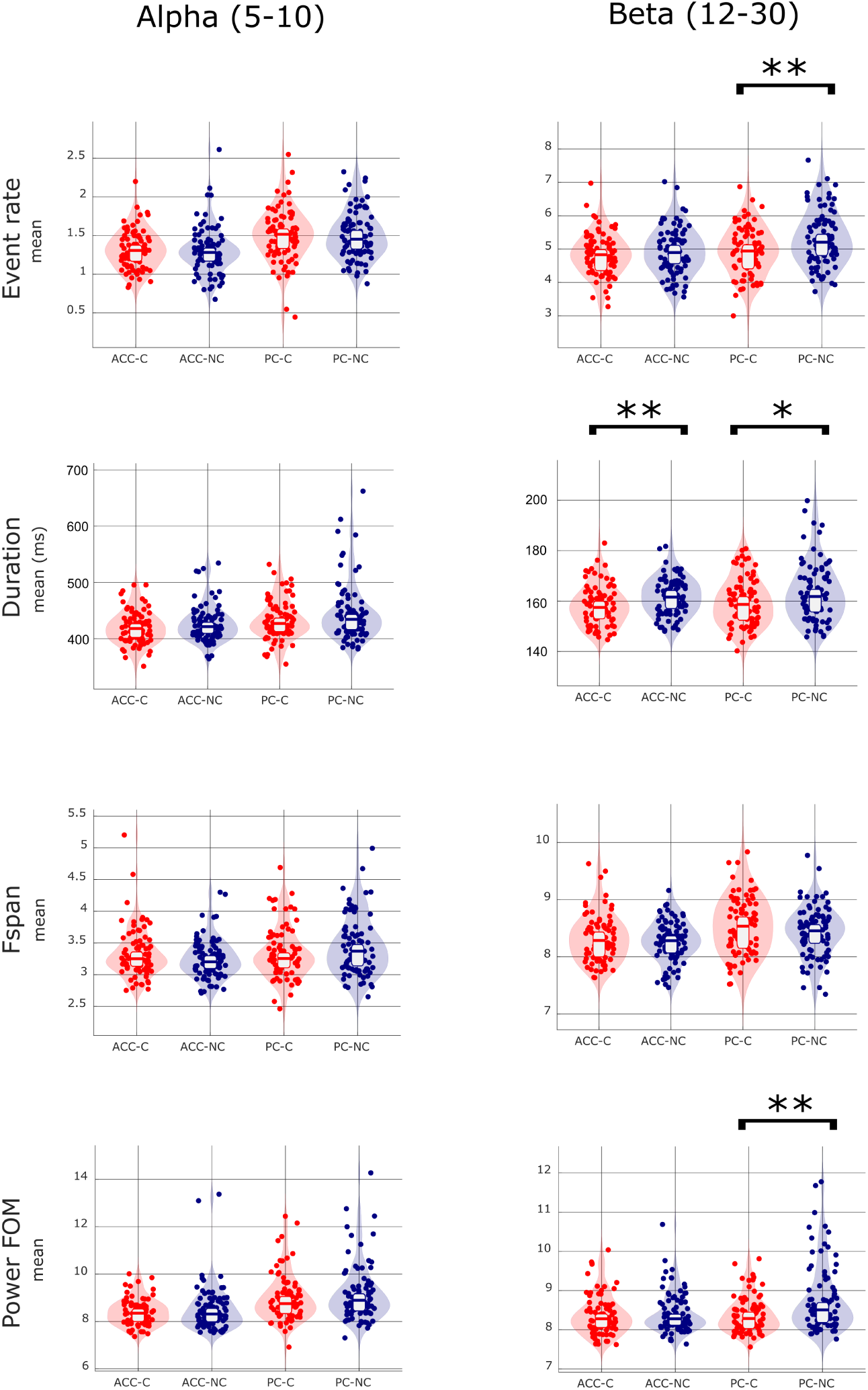
Mean and distribution of event features for AD converters (CN) and non-converters (NC) in alpha [5-10] Hz and beta [12-30] Hz frequency bands. T-test statistically significant p-values (*p <* 0.05) after BH correction are marked with the asterisk (**). Statistical tendency (*p <* 0.1) are marked with the asterisk (*).

**Table 2:** Statistical comparison of event features averaged for CONV and NOCONV groups in low-alpha [5-10] Hz and beta [12-30] Hz frequency bands and x6 FOM. Significant differences after BH correction (*p <* 0.05) are marked with the asterisk (**). Statistical tendency (*p <* 0.1) are marked with the asterisk (*).

| Freq. | ROI | Feature | CONV<br>Mean ± SD | NOCONV<br>Mean ± SD | t Stat | p | Cohen d |
| --- | --- | --- | --- | --- | --- | --- | --- |
| Low-alpha<br>[5-10] Hz | ACC | Ev.rate | 1.32 ± 0.25 | 1.30 ± 0.31 | t(165) = 0.41 | 0.343 | -0.12 |
| | | Duration | 419.6 ± 28.9 | 426.2 ± 32.3 | t(168) = -1.40 | 0.918 | 0.17 |
| | | Fspan | 3.33 ± 0.39 | 3.24 ± 0.30 | t(168) = 1.67 | 0.048 | -0.26 |
| | | Pow.FOM | 8.39 ± 0.56 | 8.51 ± 0.94 | t(144) = -1.02 | 0.844 | 0.04 |
| | PC | Ev.rate | 1.50 ± 0.34 | 1.50 ± 0.31 | t(162) = 0.13 | 0.450 | -0.08 |
| | | Duration | 432.5 ± 34.4 | 445.4 ± 51.4 | t(153) = -1.93 | 0.972 | 0.18 |
| | | Fspan | 3.33 ± 0.41 | 3.44 ± 0.45 | t(168) = -1.59 | 0.943 | 0.27 |
| | | Pow.FOM | 8.99 ± 1.00 | 9.19 ± 1.19 | t(166) = -1.17 | 0.878 | 0.16 |
| Beta<br>[12-30] Hz | ACC | Ev.rate | 4.84 ± 0.63 | 4.92 ± 0.69 | t(168) = -0.77 | 0.222 | 0.11 |
| | | Duration | 158.3 ± 8.0 | 161.7 ± 7.3 | t(164) = -2.93 | 0.002 ** | 0.48 |
| | | Fspan | 8.32 ± 0.41 | 8.26 ± 0.36 | t(161) = 1.01 | 0.844 | -0.05 |
| | | Pow.FOM | 8.38 ± 0.51 | 8.43 ± 0.51 | t(167) = -0.63 | 0.266 | 0.09 |
| | PC | Ev.rate | 4.94 ± 0.72 | 5.31 ± 0.82 | t(167) = -3.10 | 0.001 ** | 0.44 |
| | | Duration | 159.6 ± 9.4 | 163.3 ± 11.5 | t(168) = -2.25 | 0.013 * | 0.28 |
| | | Fspan | 8.54 ± 0.50 | 8.44 ± 0.43 | t(161) = 1.42 | 0.921 | -0.18 |
| | | Pow.FOM | 8.39 ± 0.46 | 8.78 ± 0.86 | t(168) = -3.69 | 0.000 ** | 0.50 |

The fact that our method allows us to explain differences in beta band, but not in alpha band, could reflect the method’s sensitivity in the alpha band. Indeed, as shown in Figure 4, in this subject sample, individual alpha event features are not a strong predictor of averaged alpha power (see also Supplementary Tables 1 & 2 for analysis of other power thresholds) and the slowing effects in the alpha band may instead be due to a combination of transient event features and/or more stationary properties of the signal.

To investigate the further predictive potential of resting-state beta event features as a biomarker for conversion from MCI to AD, we examined the association between beta event rate, duration, and maximum power and the probability of AD conversion within a 2.5-year timeframe (See Figure 6A). The probability of conversion was calculated from a logistic regression (see Methods section 2.5). Consistent with the pooled results in Figure 5, lower beta event rates, shorter duration, and reduced event power are linearly associated with an increased probability of conversion. MCI subjects with fewer than 4.92 events in four seconds of resting-state data have 1.79 (95 %CI [1.1, 3.0]) times greater odds of converting to AD (*R*^2^ = .10, *p* − *value* = .024). Those with event duration less than 159 ms have 1.06 (95 %CI [1.0, 1.1]) times greater odds of AD conversion (*R*^2^ = .06, *p* − *value* = .008), and subjects whose events have maxima power (calculated as factors of median power) less than 8.41 have 2.01 (95 %CI [1.15, 3.76]) times greater odds of AD conversion (*R*^2^ = .05, *p* − *value* = .019).

**Figure 6:**
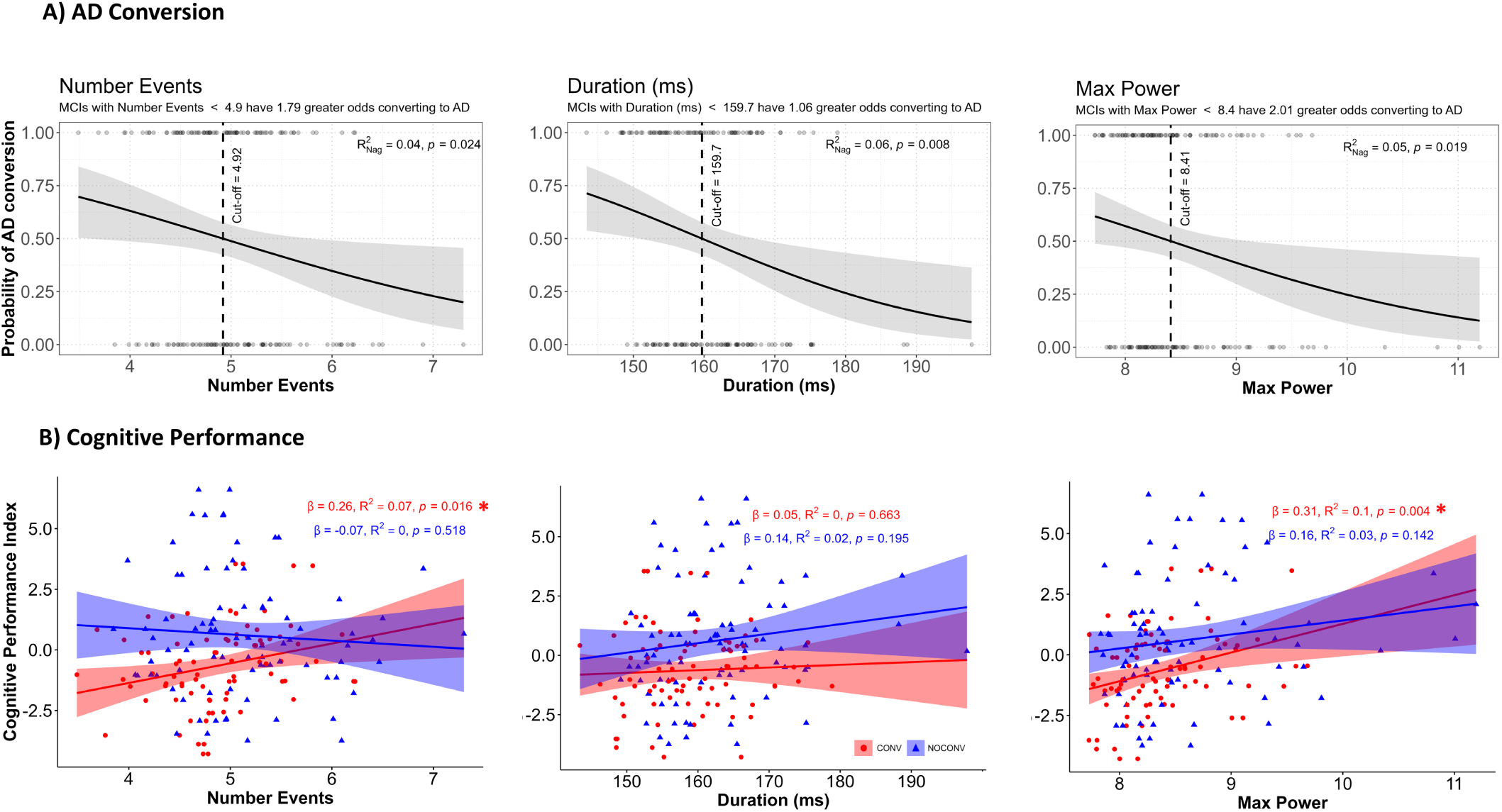
Diminished expression of beta event features in ACC and PC (number of events, duration, maxima power) is associated with greater odds of future AD conversion, and reduced cognitive performance. A) Logistic regression between beta event features and AD conversion within 2.5 years. B) Linear regression between beta event features and Cognitive Performance Index separated by CONV (red) and NOCONV (blue).

Further examination of the relationship between beta event features and cognitive performance showed that, in the CONV group, a lower rate of beta events and lower power beta events were linearly associated with greater cognitive decline (Figure 6B, lower Cognitive Performance Index values, see Methods section 2.5 *Statistical Analysis*) (*β* = .26, *R*^2^ = .07, *p* − *value* = .016 for number of events; *β* = .31, *R*^2^ = .10, *p* − *value* = .0041 for maxima power). These relationships did not emerge for duration, nor appear in NOCONVs, suggesting beta event features and particularly beta event power (*p* − *value* = 0.0041) are predictive of cognitive decline only in MCI patients that will convert to AD withing 2.5 years.

## 4 Discussion

Resting-state M/EEG signals provide a powerful non-invasive method to examine human brain physiology associated with AD (Bruña et al., 2023; Hsiao et al., 2013; Ishii et al., 2017; López et al., 2020; López-Sanz et al., 2016). Oscillatory slowing has been associated with AD conversion based on PSD analysis that relies on signal averaging. Our study shows for the first time that in non-averaged data, resting state alpha and beta oscillations from ACC and PC are composed of transient high-power events. To explore how transient high power events relate to findings of slowing in PSD, we studied their properties in a sample of individuals diagnosed as MCI and characterized novel features of transient high-power events that distinguish patients who later convert or do not convert to AD (i.e., CONV and NOCONV, respectively). Our analysis reveals a consistent pattern of a lower number of transient 12-30 Hz beta events, duration and power for CONV compared to NOCONV in PC, with the duration effect also occurring in AC. This diminished beta event expression in ACC and PC was associated with increased odds of future AD conversion and decreased cognitive performance in the CONV group. It is well known that PC and ACC are part of the default mode network, which has decreased metabolism in the early stages of the disease (Greicius et al., 2004) and it is closely involved with episodic memory processing (Liu et al., 2022). Our finding of reduced event-activation in CONV is predominantly found in precuneus, a ROI typically associated with deposition of amyloid-*β* in the early stages of the AD continuum (Forsberg et al., 2008).

Overall, our results lay the foundation for further examination of beta event features as a novel biomarker for early AD diagnosis and a possible neurobiological measures of the effectiveness of preventative interventions. The more fine-grained description of slowing in unaveraged MEG data may also bring us closer to understanding the underlying neural mechanisms, and a more direct link to hyperexcitability as observed in animal models (Maestú et al., 2021; Zott et al., 2019), ultimately guiding new therapeutics in humans.

### 4.1 Consistency with prior studies examining beta, ageing and cognitive control

Reductions in beta event expression yielded a lower PSD in the beta band [12-30] Hz averaged across trials, giving further insight into the underpinning of oscillatory slowing in AD (Bruña et al., 2023; Dauwels et al., 2011; Jelic et al., 2000). This trend is also consistent with a critical shift in beta activity at approximately 60 years of age in healthy patients (Brady et al., 2020; Power & Bardouille, 2021).

Following this inflection point, resting-state relative source power, as well as beta event characteristics such as event rate, peak frequency, duration, peak power, etc., progressively begin to decline with age (Brady & Bardouille, 2022). Other studies have shown that averaged M/EEG spectral activity in the beta frequency range (13-30) Hz is a more powerful predictor of MCI-to-AD conversion than activity in other frequency bands, including in the slow alpha frequency range ((Gaubert et al., 2024; Poil et al., 2013). Thus, transient beta event features may be key factors in delineating differences and tracking the neurophysiology of healthy and pathological ageing.

We found that reductions in beta event features were associated with global cognitive decline, as measured by an index based on a battery of neuropsychological tests. However, this effect is only significant in the CONV group. MCIs who convert to AD 2.5 years later seem to be more reliant on beta event expression for cognitive robustness.

Several studies have shown that beta events throughout the cortex are a signature of inhibitory control. In sensory cortex, beta event expression can be manipulated with attention (increasing in non-attentive states), and an increase in beta event rates is associated with a decrease in perceptual salience (Shin et al., 2017), a process predicted to be mediated by an increase in inhibitory neuron activity (Law et al., 2022). In motor cortex, increased beta event expression is associated with inhibited motor control and Parkinson’s disease (Yu et al., 2021), and in frontal cortex beta events are a signature of stopping of movement and long-term memory retrieval (Schmidt et al., 2019; Wessel, 2020). Conversely, reduction in beta event rates in frontal cortex has been explicitly linked to encoding and decoding in working memory processes, where they have been suggested as a mechanism for volitional control and memory content reactivation (Lundqvist et al., 2016, 2018; Spitzer & Haegens, 2017). Likewise, decreased posterior parietal beta oscillatory activity predicts episodic memory formation and retrieval (Griffiths, Martín-Buro, Staresina, Hanslmayr, & Staudigl, 2021; Nyhus, 2018), and is correlated with enhanced memory performance (Griffiths, Martín-Buro, Staresina, & Hanslmayr, 2021). Transcranial stimulation of the prefrontal cortex at beta (18 Hz) has been found to induce memory encoding impairments (Hanslmayr et al., 2014), and during posterior parietal cortex stimulation, lower prestimulus beta power predicts higher phosphene ratings, reflecting increased neural excitability (Samaha et al., 2017).

The consistent relationship between beta event expression and inhibitory control in these myriad studies (Lundqvist et al., 2024) suggests that the ability to modulate beta events according to task demands is necessary for optimal function, and that the diminished resting state beta event expression and associated cognitive decline observed in the PC in CONV in our study may be directly related to lack of inhibitory cognitive control.

### 4.2 Why are differences in alpha bursts not present?

Despite the observation of significant across group differences in alpha band in the average PSD (Figure 2), we did not find a significant relationship between transient [5-10] Hz alpha event features and averaged power, nor did we find a difference in event features between MCI patients who will convert to AD and those who will not, which is consistent with previous longitudinal studies of AD progression (Gaubert et al., 2024; Poil et al., 2013). This was true for several event detection thresholds (see Supplementary Figure 2 and Supplementary Tables 1 and 2).

Since we did not find a relationship between spectral events in the lower 5-10 Hz alpha band and averaged alpha power in our sample, we did not examine the relationship to cognitive performance. In terms of components of slowing, other studies have found a relationship between bursts of slow activity (1-6 Hz) age and cognition in healthy adults, where older subjects and lower cognitive performance participants exhibited longer and slower events (Power et al., 2024).

The lack of alpha event effects in our sample could reflect the sensitivity limitations of the spectral event detection method or indicate that, although the alpha band contains transient components, the differences between the two groups involve a combination of transient feature or more stationary features than transient ones. A slowed occipital alpha rhythm is a common biomarker of several neurological or psychiatric disorders (Hughes & Crunelli, 2005; Samson-Dollfus et al., 1997). According to the “thalamo-cortical (TC) hypothesis” (Klimesch et al., 2007; S. M. Sherman, 2001) slow cortical rhythms could be generated by TC cells in tonic, single spike, firing, or in arhythmic bursting mode. Hugues (2005) argued that a slowing effect when shifting from alpha to theta waves could arise from the hyperpolarization of the thalamo-cortical neuron population, resulting in a deceleration of high-threshold bursting (HT) in individual cells, which could translate to a shift from bursty alpha to more continuous slower theta range oscillations. Thus, in this cognitively impaired population, while we still detect some alpha bursting, Alzheimer’s disease-related mechanisms may be disrupting thalamo-cortical connections (Eustache et al., 2016) and slowing the thalamic bursting activity, making it more stationary.

In the early stages of AD continuum, even before a mild cognitive impairment diagnosis, alpha disruption is a marker of cognitive decline (Babiloni et al., 2010; Bruña et al., 2023; Huang et al., 2000; Ĺopez et al., 2020; López-Sanz et al., 2016). Still, when patients reach a more advanced stage of the disease, as it is mild cognitive impairment, beta band is more discriminant between AD converters and non-converters (Gaubert et al., 2024; Poil et al., 2013; Pusil, Dimitriadis, et al., 2019). Consequently, when measured in MCIs the spectral events method may not be sensitive to this transient-to-stationary shift in the alpha [5-10] Hz frequency band but can detect differences in the beta [12-30] Hz band.

### 4.3 A mechanistic link between reduced beta event expression and hyperexcitability in CONV

Animal studies have suggested A*β*-induced changes in E/I balance is a mechanism for hyperexcitability and cognitive decline in Alzheimer’s disease (Maestú et al., 2021). This notion has been supported by several computational neural modeling frameworks examining causal links between disease processes and increased neuron firing rates (Alexandersen et al., 2023; Cabral et al., 2014; de Haan et al., 2017; Hutt et al., 2023; Nakagawa et al., 2014; Stefanovski et al., 2019; Zimmermann et al., 2018), albeit without explicit consideration of the biophysical generators of MEG current sources or of beta frequency oscillations.

Modeling work by our group specifically designed to interpret the detailed cell and circuit origin of localized MEG (and EEG) current source signals (Neymotin et al., 2020) provides a hypothesized direct mechanistic link between beta event expression and hyperexcitability in CONV. Specifically, our prior modeling and cross-species empirical studies suggested neocortical beta events are generated by bursts of exogenous thalamocortical drive that targets excitatory synapses on the proximal and distal dendrites of pyramidal neurons in deep and superficial neocortical layers, such that the distal drive is stronger and last a beta period (i.e. ≈ 50ms) (Jones et al., 2009; M. A. Sherman et al., 2016). This thalamic burst drive induces current flow in pyramidal neuron dendrites that generates MEG beta event waveform characteristics consistent with those observed experimentally in sensory, motor and frontal cortices (Bonaiuto et al., 2021; M. A. Sherman et al., 2016), and when occurring rhythmically can produce multiple beta cycles and/or a complex of alpha and beta activity (Jones et al., 2009). Follow-up studies predicted further that the thalamic drive inducing a beta event also activates inhibitory neurons in supragranular layers, providing a causal mechanism for beta-associated inhibitory control (Law et al., 2022; Shin et al., 2017). Together with our current findings that CONV have reduced beta expression (namely lower event rates, power and duration), these prior studies suggest CONV have a reduction in thalamocortical burst drive to cortex, which in turn recruits less cortical inhibitory neuron activity leading to hyperexcitability.

Such a decrease in inhibitory activity could contribute to the observation of diminished Gabaeric terminals on cortical neurons near amyloid plaques, in addition to the toxic effects of amyloid oligomers on inhibitory terminals (Alexandersen et al., 2023; Garcia-Marin et al., 2009), particularly in PC which exhibits A*β* deposition in the early stages of AD. The predicted reduction in thalamic bursting is also synergistic with studies showing thalamic atrophy and reduced inhibitory thalamic tone (Abuhassan et al., 2014; Forno et al., 2023), as inhibition from the thalamic reticular nucleus is known to be a driver of rebound bursting mechanisms in thalamic relay cells (Destexhe & Sejnowski, 2002). Moreover, decreases in thalamocortical drive are consistent with theories of thalamocortical dysrhythmia (Llińas et al., 1999) and could contribute to loss of cortical signal complexity and biophysical heterogeneity (Szul et al., 2023) occurring with the disease and thought to be an important homeostatic control mechanisms capable of bolstering the network’s resilience to perturbations, such as the toxic effects of amyloid (Dauwels et al., 2011; Hutt et al., 2023).

## Data Availability

All data produced in the present study are available upon reasonable request to the authors

## Supplementary Material

### “Mirror” events

The MEG activity of medial regions is characterized by the presence of synchronized, contralateral, spurious mirror events (see TFRs in Figure S1). This phenomenon is caused by the limited resolution of MEG/volume conduction. When using the MNE dipole-constrained source reconstruction method, these duplicated events are noticeable because they are characterized by a mutual inversion of the time series. We performed a removal step of duplicated mirror events: when two detected events shared the same (but contralateral) ROI, epoch, time and frequency, we discarded the one with lower amplitude.

**Figure 1:**
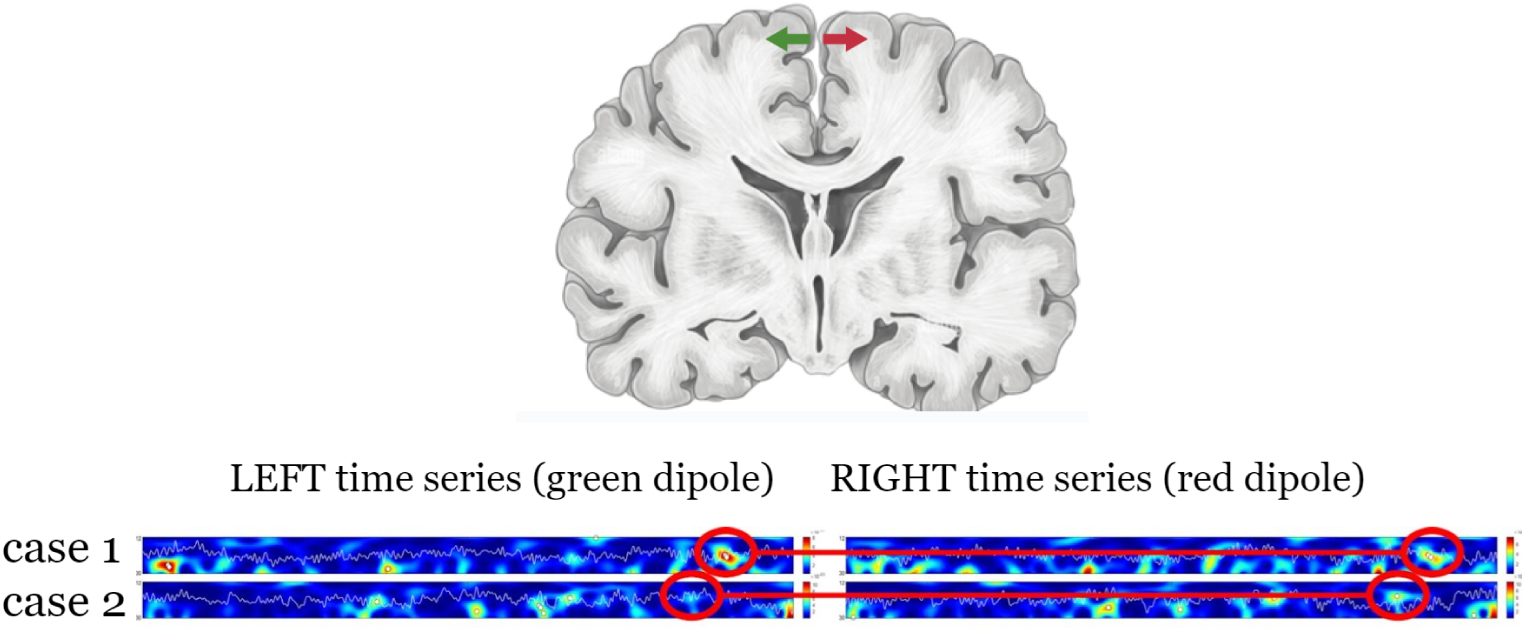
Example of mirror event in left ACC and right ACC due to spatial smearing in source reconstruction. The events considered at the end of the detection process were the left one in the case1, and the right one in the case2, due to their amplitudes.

### Event threshold analysis

**Figure 2:**
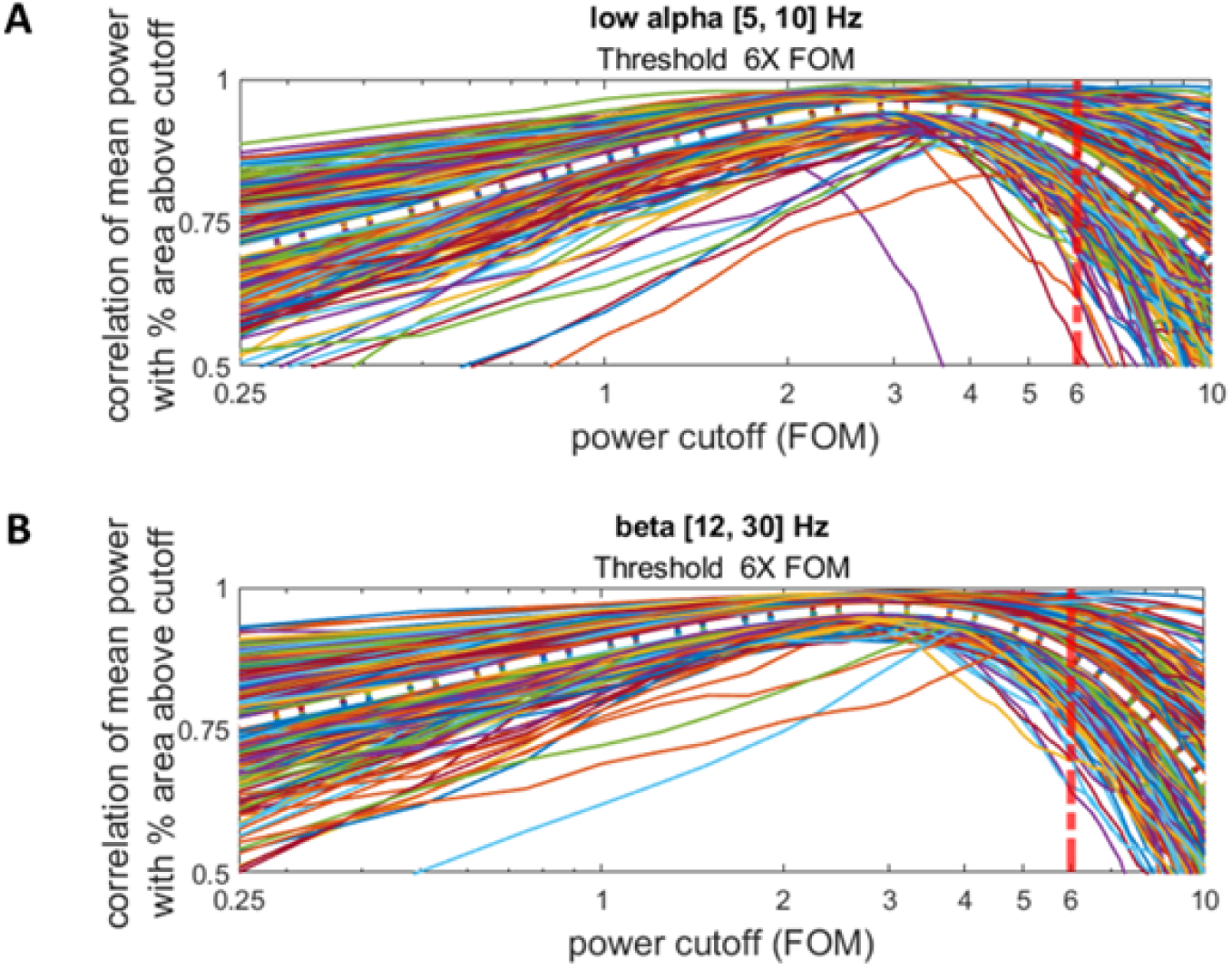
Average relationship between power cutoff FOM and correlation of mean power with percentage of area above cutoff. a) Analysis over events selected in alpha frequency [5-10] Hz and visualisation of 6X threshold (red line). b) Analysis over events selected in beta frequency [12-30] Hz and visualisation of 6X threshold (red line).

**Table 1:** Statistical comparison for x4 FOM of event features mean averaged for CONV and NOCONV groups in low-alpha [5-10] Hz and beta [12-30] Hz frequency bands. Significant differences (*p <* 0.05) are marked with the asterisk (**). Statistical tendency (*p <* 0.1) is marked with the asterisk (*).

| Freq. | ROI | Feature | t-Stat | p-value | BH adjusted p-value | Cohen d |
| --- | --- | --- | --- | --- | --- | --- |
| Low alpha [5-10] Hz | ACC | Ev. Rate | 1,705 | 0,045 | 0,18 | -0,226 |
|  |  | Duration | -2,084 | 0,981 | 0,993 | 0,258 |
|  |  | Fspan | 1,484 | 0,07 | 0,203 | -0,126 |
|  |  | Pow.FOM | -1,11 | 0,866 | 0,993 | 0,078 |
|  | PC | Ev. Rate | 1,113 | 0,134 | 0,267 | -0,188 |
|  |  | Duration | -2,507 | 0,993 | 0,993 | 0,376 |
|  |  | Fspan | -1,453 | 0,926 | 0,993 | 0,267 |
|  |  | Pow.FOM | -1,221 | 0,888 | 0,993 | 0,16 |
| Beta [12-30] Hz | ACC | Ev. Rate | 1,716 | 0,956 | 0,993 | -0,217 |
|  |  | Duration | -2,67 | 0,004 | 0,033 ** | 0,383 |
|  |  | Fspan | 1,241 | 0,892 | 0,993 | -0,086 |
|  |  | Pow.FOM | -1,438 | 0,076 | 0,203 | 0,224 |
|  | PC | Ev. Rate | -1,183 | 0,119 | 0,267 | 0,194 |
|  |  | Duration | -2,37 | 0,009 | 0,05 * | 0,336 |
|  |  | Fspan | 2,213 | 0,986 | 0,993 | -0,358 |
|  |  | Pow.FOM | -3,744 | 0 | 0,002 ** | 0,561 |

**Table 2:** Statistical comparison for x8 FOM of event features mean averaged for CONV and NOCONV groups in low-alpha [5-10] Hz and beta [12-30] Hz frequency bands. Significant differences (*p <* 0.05) are marked with the asterisk (**). Statistical tendency (*p <* 0.1) is marked with the asterisk (*).

| Freq. | ROI | Feature | t-Stat | p-value | BH adjusted p-value | Cohen d |
| --- | --- | --- | --- | --- | --- | --- |
| Low alpha [5-10] Hz | ACC | Ev. Rate | -0,497 | 0,69 | 0,91 | -0,077 |
|  |  | Duration | 0,679 | 0,249 | 0,639 | -0,247 |
|  |  | Fspan | 0,082 | 0,467 | 0,91 | 0,037 |
|  |  | Pow.FOM | -0,33 | 0,629 | 0,91 | -0,029 |
|  | PC | Ev. Rate | -0,585 | 0,72 | 0,91 | 0,078 |
|  |  | Duration | -0,959 | 0,831 | 0,91 | 0,047 |
|  |  | Fspan | -1,052 | 0,853 | 0,91 | 0,164 |
|  |  | Pow.FOM | -0,804 | 0,789 | 0,91 | 0,162 |
| Beta [12-30] Hz | ACC | Ev. Rate | -0,585 | 0,28 | 0,639 | 0,095 |
|  |  | Duration | -1,831 | 0,034 | 0,138 | 0,287 |
|  |  | Fspan | 0,295 | 0,616 | 0,91 | -0,007 |
|  |  | Pow.FOM | -0,973 | 0,166 | 0,531 | 0,083 |
|  | PC | Ev. Rate | -3,446 | 0 | 0,003 ** | 0,466 |
|  |  | Duration | -2,221 | 0,014 | 0,074 * | 0,317 |
|  |  | Fspan | 1,721 | 0,956 | 0,956 | -0,22 |
|  |  | Pow.FOM | -3,602 | 0 | 0,003 ** | 0,498 |

**Figure 3:**
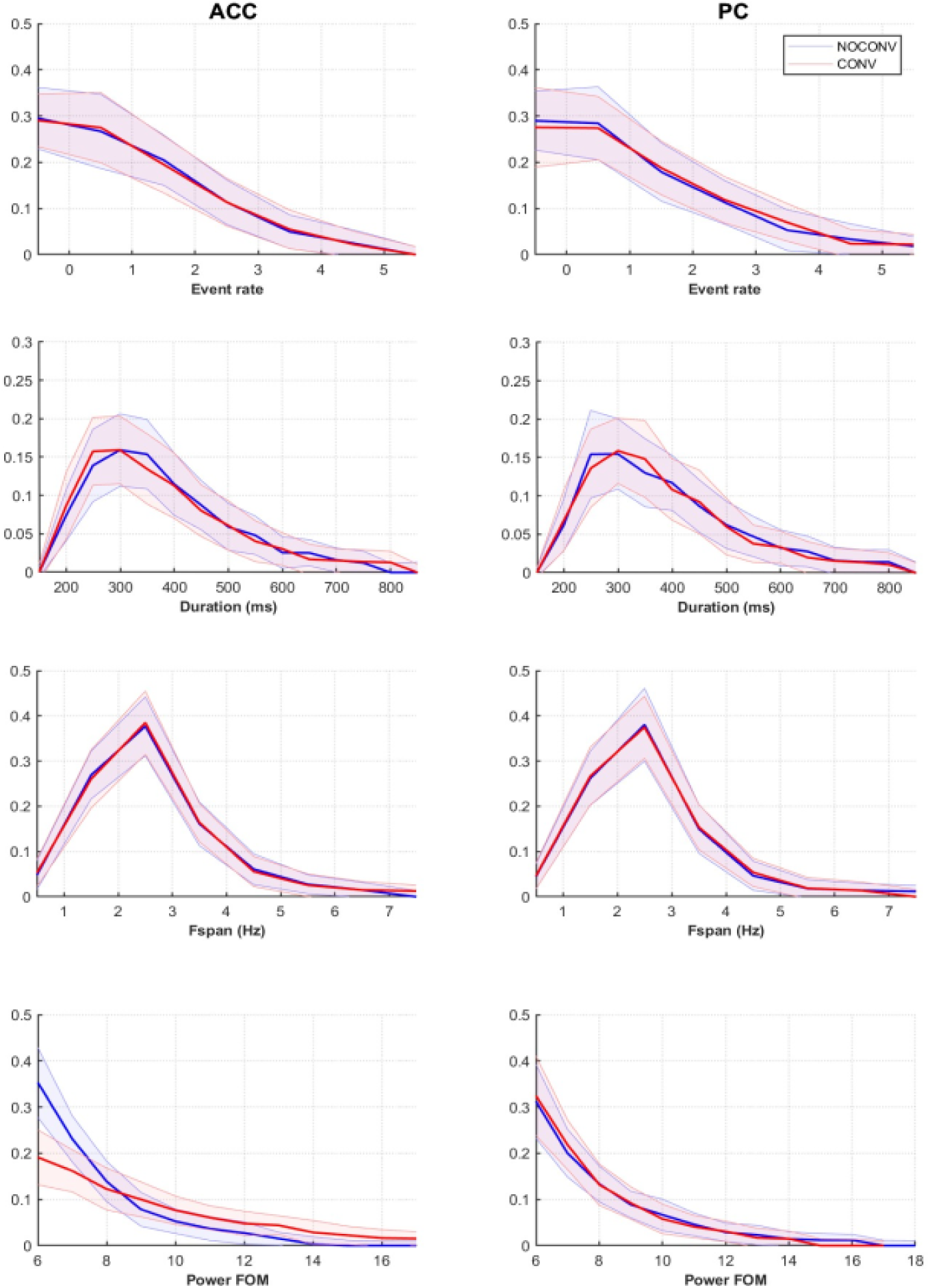
Probability density plots for each event features in low-alpha [5-10] Hz band for 4 seconds trials.

**Figure 4:**
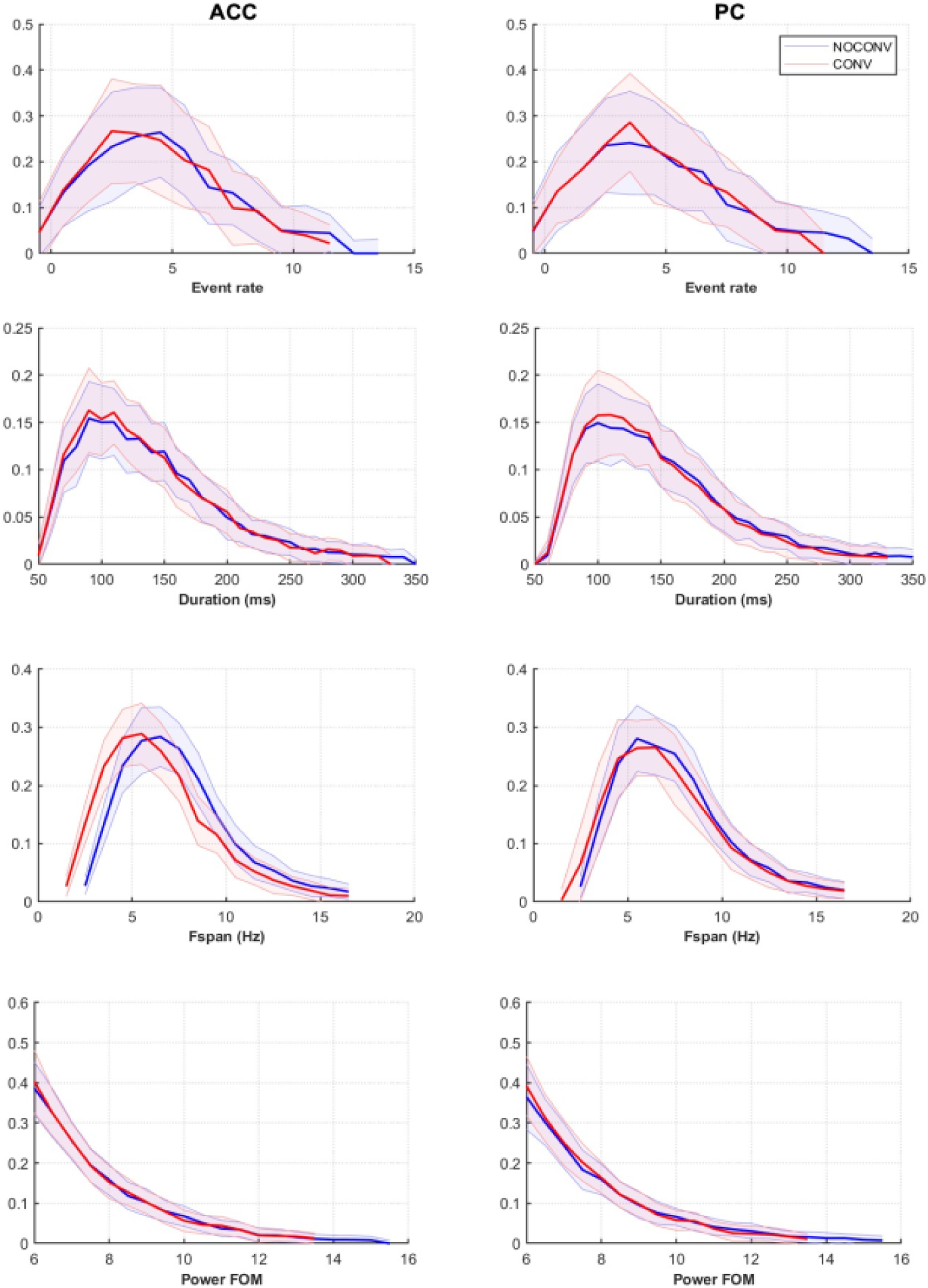
Probability density plots for each event features in beta [12-30] Hz band for 4 seconds trials.

## Data and Code Availability

Code to perform analyses is available at: https://github.com/jonescompneurolab/SpectralEvents

Data will be made available on request.

## Author Contributions

DSB, GL, RB – methodology, software, analysis, visualization, original draft

DZ, JC, BC, EP, ME, RB, FM, SJ – conceptualization, methodology, review and editing.

